# COVID-19 Outcomes and Genomic characterization of SARS-CoV-2 isolated from Veterans in New England States

**DOI:** 10.1101/2021.04.27.21256222

**Authors:** Megan Lee, Ya Haddy Sallah, Mary E. Petrone, Matthew Ringer, Danielle Cosentino, Chantal B.F. Vogels, Joseph R. Fauver, Tara Alpert, Nathan D. Grubaugh, Shaili Gupta

## Abstract

In an older cohort of veterans with a high comorbidity burden, age was the largest predictor of hospitalization, peak disease severity, and mortality. Most infections in six New England states until September, 2020, were from SARS-CoV-2 B.1 lineage, dominated by spike protein D614G substitution in 97.3% of samples.

## Background

Disease severity and outcomes of Coronavirus Disease 2019 (COVID-19) caused by Severe Acute Respiratory Syndrome Coronavirus-2 (SARS-CoV-2) vary among individuals who become infected, with several factors that have been suggested as predictors of mortality, including Charlson comorbidity index score, age, and body mass index (BMI) [2-6]. Virologic characteristics have also been suggested to impact the severity of disease, and concern has been raised about several variants being more transmissible or more lethal [7,8,9,10]. A dynamic nomenclature system (Pangolin) was developed to classify SARS-CoV-2 and identify lineages and mutations that could impact infectivity and virulence [11]. Understanding viral epidemiology and a regional evaluation of viral variants, with their respective clinical correlations, is important to provide a full understanding of the disease.

Given the high variability and conflicting data in predicting who will have poor outcomes, assessment of specific populations is necessary to give providers the best clinical picture on their patients. Clinical outcomes of veterans with COVID-19 in New England and respective SARS-CoV-2 genomics have not been described. We aimed to (1) describe patient characteristics, comorbidities, and disease factors that impact patient outcomes and (2) present data on the genomic composition of the SARS-CoV-2 infecting these patients.

## Study Design

Our cohort was composed of all veterans receiving care at Veterans Administration (VA) healthcare centers in six New England states (Connecticut, Massachusetts, Maine, New Hampshire, Rhode Island, Vermont) who tested positive for COVID-19 from April 8, 2020, to September 16, 2020. Inclusion criteria included patients with accessible chart records and a diagnosis of COVID-19 based on one of three tests: Xpert^®^ Xpress SARS-CoV-2 (Cepheid), Simplexa ^®^ COVID-19 Direct Kit (DiaSorin), and Roche cobas^®^ 6800 system. We manually reviewed charts and recorded demographics [age, gender, race, body-mass index (BMI), long-term care facility (LTC) status, and state of residence when diagnosed with COVID-19]. Comorbidities recorded were immunosuppression, dementia, diabetes mellitus, chronic kidney disease stage ≥ 3, chronic liver disease, coronary artery disease (CAD), heart failure, atrial fibrillation, chronic obstructive pulmonary disease, asthma, and active tobacco use. Clinical database and Viral RNA repositories were created and approved by VA Connecticut Institutional Review Board. Whole genome sequencing was conducted on SARS-CoV-2 isolates with a cycle threshold (Ct) value <36, and provided near-complete or complete genome results where Ct value was <30.

We recorded hospitalization status, mortality, and oxygen (O2)-requirement within 24 hours of admission. We divided patients based on their peak disease severity into five categories depending on oxygenation requirements: 1) no O2-requirement, 2) 1-3 liters (L) by nasal cannula (NC), 3) 4-6 L NC, 4) >6 L O2 or non-invasive positive pressure ventilation, and 5) mechanical ventilation. We used STATA v16 for univariate and multivariate logistic regressions to predict the outcomes of interest.

We sequenced whole virus genomes (≥20x coverage depth across ≥70% of the genome) using Illumina (n = 238) and Nanopore (n = 61) platforms. Using BWA-MEM version 0.7.15, we aligned reads to the Wuhan-Hu-1 reference genomes (GenBank MN908937.3). With iVar v1.2.1, we trimmed sequencing adaptors and primer sequences and called bases by simple majority (>50% frequency) at each site to generate consensus genomes. An ambiguous N was used when < 10 reads were present at a site. We aligned consensus genomes with MAFFT [12], and masked problematic sites [13]. We built a phylogenetic tree with IQTree [14] using an HKY substitution model and 1000 bootstraps, visualized it with Python module baltic v0.1.5, and assigned lineages with Pangolin [11].

## Results

Of 476 veterans in six New England states with confirmed SARS-CoV-2 during the study period, 274 had complete and accessible charts. Of 274 veterans, 92.7% were men, 83.2% White, and mean age was 63 years (standard deviation: 17.6 years) and over a third resided in LTC (n=92) (Supplementary Table 1). The most common comorbidities were CAD (27%), diabetes (25%), and tobacco use (23%).

Notably, 11.7% patients required O2 above their baseline home O2-requirement within 24 hours of admission, and 20.8% of all patients required O2 support at some point during hospitalization. In terms of peak severity, 78.5% required only room air, 10.9% required 1-3 L O2, 4.0% required 4-6 L O2, 3.6% required >6L O2 or NIPPV, and 2.2% were intubated. The hospitalization rate was 28.8% (Supplementary Figure 1), and overall mortality rate was 10.6% (Supplementary Figure 2).

Univariate regression analysis results are reported in Table 1. In multivariate regression, significant predictors of hospitalization were age (OR: 1.05) and non-White race (OR: 2.39) (Table 3). Peak severity also varied by age (OR: 1.07) and O2 requirement on admission (OR: 45.7). Mortality was predicted by age (OR: 1.06), dementia (OR:3.44), and O2 requirement on admission (OR: 6.74). In other words, for every year increase in age, the odds of hospitalization increased by 5%, peak severity increased by 7%, and mortality increased by 6%.

**Table 1.**
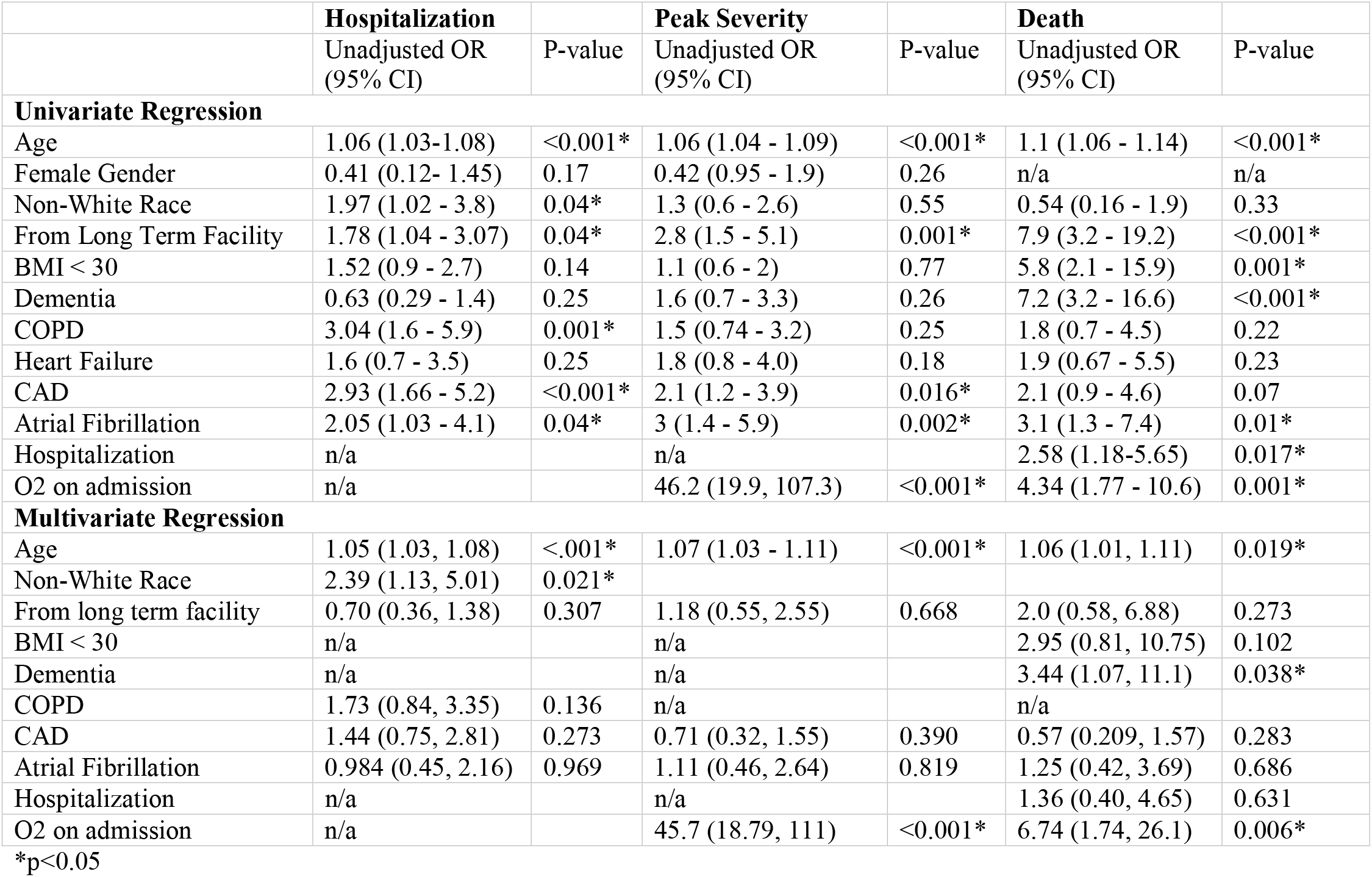
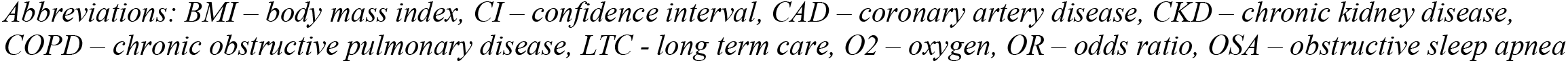
Univariate and multivariate regression analysis of factors that predict hospitalization, peak severity, and death

For the genomic characteristics, amongst the 426 patients, 299 patients’ genomes had adequate coverage for analysis. We found the majority of our specimens (51.5%) were from SARS-CoV-2 lineage B.1, or a sub-lineage of B.1 (e.g. B.1.302, B.1.303, B.1.356; 137/299, 45.8%), all of which are defined by D614G substitution (Figure 1, Supplementary Table 2). Only 2.4% were from the lineage A that lack the D614G mutation. There were 41 SARS-CoV-2 lineages detected in our cohort, and we did not have the power to test for clinical correlates. Our sequencing data does inform us that the outcomes presented in this VA cohort are dominated by the impacts of the B lineage D614G mutation.

**Figure 1.**
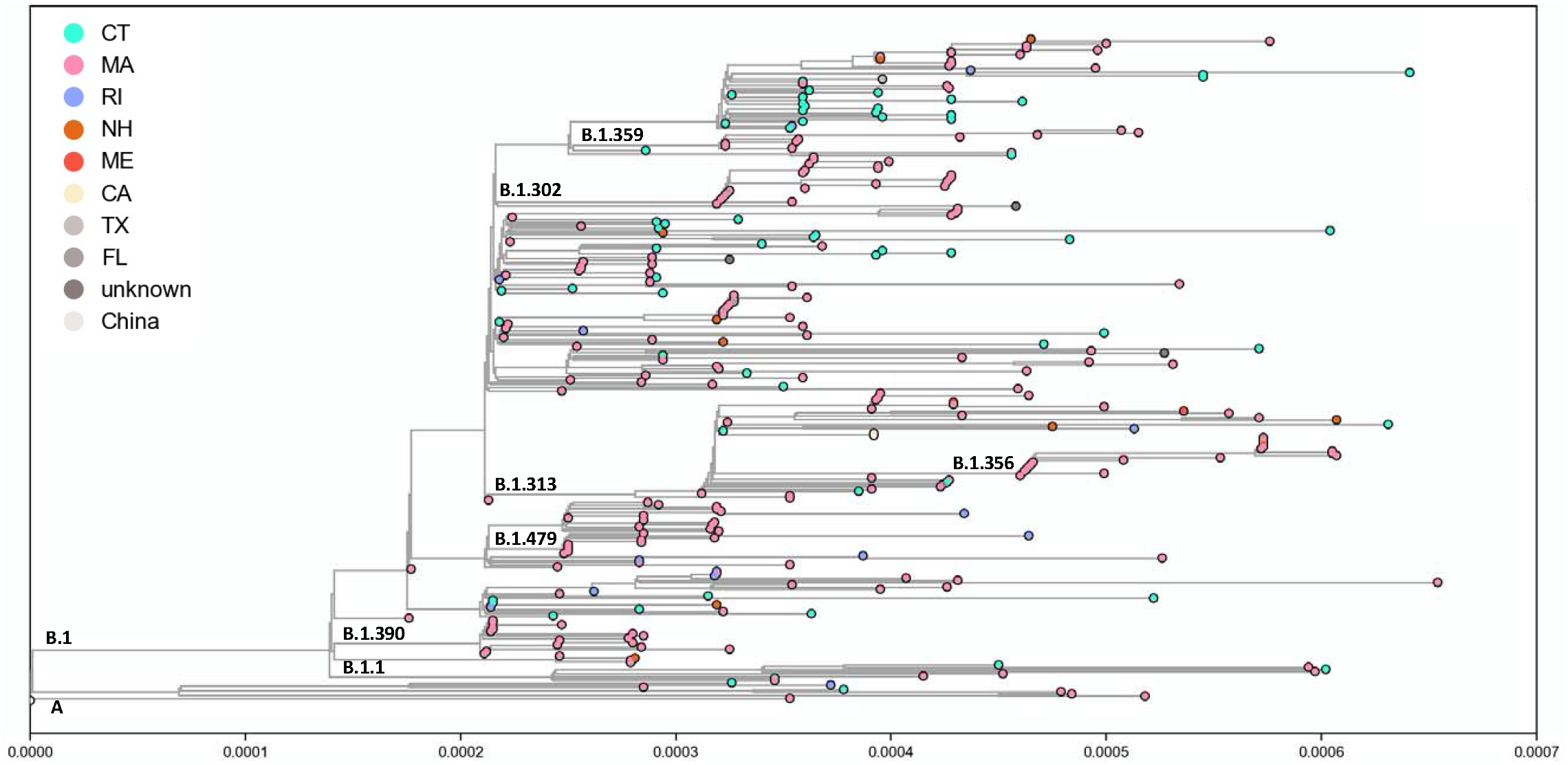
Maximum Likelihood tree of genomes

## Discussion

Our study found that in an older cohort of veterans with a high comorbidity burden, age significantly associated with risk of hospitalization, peak disease severity, and mortality. The CDC provides a list of chronic medical conditions that predispose individuals to severe illness from SARS-CoV-2 infection [15], but >75% of United States adults fall under a high-risk category [16]. Veterans are a unique cohort because of advanced age on average, and more comorbidities. Understanding clinical factors that impact outcomes in veterans will help clinicians risk-stratify patients with similar demographic profiles.

Many COVID-19 studies have found age to be a predictor of worse outcomes [4,17-20]. In our study, age was a significant predictor for all of our outcomes and was a confounder for other variables. Interestingly, LTC status predicted all three of our outcomes on univariate analysis, but not on multivariate analyses. Earlier in the COVID-19 pandemic, residents of nursing homes had higher rates of infection as well as severe illness and mortality [21]. Our study shows that among veterans in LTC facility, disease outcomes were not impacted by their residence status.

Concurrent work from our group suggests that O2-requirement within 24 hours of admission predicts poor outcomes in veterans, which has helped inform the triage guidelines at our healthcare system. This is an important finding because other ways of determining oxygenation status can frequently change, and thus become difficult for clinicians to use in practice [22].

Our study supports data from previous reports that non-White patients are at increased risk of hospitalization but have similar peak severity and mortality outcomes [23-26]. Many studies have shown that minorities often have delays in seeking care, causing higher risk of hospitalization when they do seek care [27,28]. This may explain the outcomes in our study. It is critical to continue ongoing efforts to combat medical inequities and target prevention efforts and education to communities and racial groups most affected by COVID-19.

After adjusting for age and other comorbidities, we found that patients with dementia had a higher risk of death. This is similar to other studies on patients with COVID-19 and dementia [18, 29-31]. This may be explained by a host of biological factors, but it emphasizes the importance of extra care and monitoring required when approaching a patient with dementia.

Limitations of this work include the smaller sample size. Furthermore, our study is specific to veterans, which is a largely male and older cohort, and results may not therefore be generalizable. The time period of this study was prior to established medical therapies for COVID-19 and our outcomes reported are likely worse than expected today. Strengths of our study include its comprehensive scope, wide geographic range, manual chart review allowing for the capturing of all comorbidities and oxygenation parameters that may not be available otherwise in a database, and multivariate analysis of many potential risk factors.

## Conclusion

Our study found that in an older cohort of veterans from the six New England states with a high comorbidity burden, age was the single strongest predictor of hospitalization, peak severity, and mortality. Non-white veterans were more likely to be hospitalized, and patients who required oxygen on admission were more likely to have severe disease and higher rates of mortality. Furthermore, patients with dementia were more likely to die. Multiple genomic variants of SARS-CoV-2 were distributed in patients in New England early in the COVID-19 era, mostly from a B.1 sub-lineage with the spike D614G mutation.

## Data Availability

Data is available upon request

## Acknowledgements

N.D.G. is a paid consultant of Tempus labs for infectious disease genomics. The other authors report no conflicts of interest.

---

**Supplemental Table 1.** Patient Characteristics (n=274)

| **Demographics** |  | **Comorbidities** |  |
| --- | --- | --- | --- |
| Age (years) | 62.9+17.6 | Immunosuppressed | 10 (4) |
| Gender |  | Dementia | 42 (15) |
| Male | 254 (93) | Diabetes | 68 (25) |
| Female | 20 (7) | CKD 3 | 18 (7) |
| Race |  | Chronic liver disease | 32 (12) |
| White | 228 (83) | Chronic heart disease | 107 (39) |
| Non-white | 46 (17) | CAD | 74 (27) |
| BMI > 30 | 110 (40) | Heart failure | 29 (11) |
| From LTC | 92 (34) | Atrial fibrillation | 40 (15) |
| State |  | Chronic Lung disease | 97 (35) |
| Connecticut | 89 (32) | COPD | 44 (16) |
| Massachusetts | 150 (55) | Asthma | 19 (7) |
| Maine | 4 (1) | OSA | 55 (20) |
| New Hampshire | 9 (3) | Tobacco Use | 62 (23) |
| Rhode Island | 20 (7) |  |  |
| Vermont | 2 (1) |  |  |

*Results are presented as numbers (percentages).*
*Abbreviations: LTC - long term care, BMI – body mass index, CKD – chronic kidney disease, CAD – coronary artery disease, COPD – chronic obstructive pulmonary disease, OSA – obstructive sleep apnea*

**Supplemental Table 2.** Lineages of genomes

| Lineage | Count | Percent |
| --- | --- | --- |
| A.1 | 3 | 1.0% |
| A.2 | 2 | 0.7% |
| A.3 | 2 | 0.7% |
| B | 1 | 0.3% |
| B.1 | 154 | 51.5% |
| B.1.1.10 | 1 | 0.3% |
| B.1.1.113 | 1 | 0.3% |
| B.1.1.128 | 1 | 0.3% |
| B.1.1.163 | 1 | 0.3% |
| B.1.1.192 | 1 | 0.3% |
| B.1.1.225 | 1 | 0.3% |
| B.1.1.307 | 1 | 0.3% |
| B.1.104 | 4 | 1.3% |
| B.1.108 | 1 | 0.3% |
| B.1.110.3 | 1 | 0.3% |
| B.1.2 | 3 | 1.0% |
| B.1.243 | 1 | 0.3% |
| B.1.268 | 1 | 0.3% |
| B.1.280 | 2 | 0.7% |
| B.1.302 | 26 | 8.7% |
| B.1.313 | 19 | 6.4% |
| B.1.319 | 1 | 0.3% |
| B.1.320 | 1 | 0.3% |
| B.1.331 | 1 | 0.3% |
| B.1.356 | 18 | 6.0% |
| B.1.359 | 8 | 2.7% |
| B.1.36 | 1 | 0.3% |
| B.1.369 | 3 | 1.0% |
| B.1.382 | 1 | 0.3% |
| B.1.385 | 1 | 0.3% |
| B.1.390 | 16 | 5.4% |
| B.1.403 | 1 | 0.3% |
| B.1.413 | 1 | 0.3% |
| B.1.448 | 2 | 0.7% |
| B.1.448 | 1 | 0.3% |
| B.1.479 | 10 | 3.3% |
| B.1.509 | 2 | 0.7% |
| B.1.517 | 1 | 0.3% |
| B.1.521 | 1 | 0.3% |
| B.35 | 1 | 0.3% |
| N.1 | 1 | 0.3% |

**Supplemental Figure 1.**
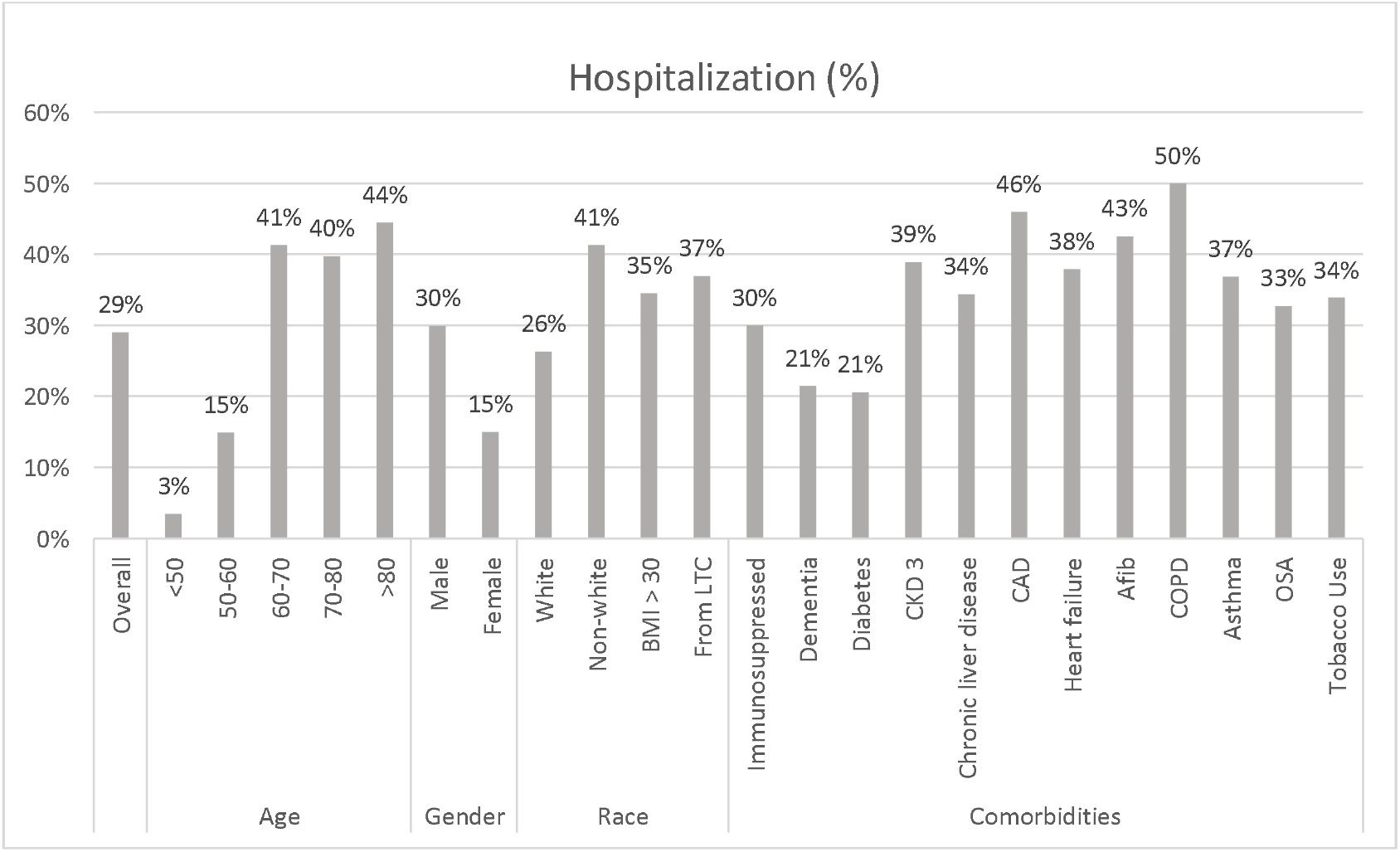
Percent of patients hospitalized, based on patient demographics and comorbidities

**Supplemental Figure 2.**
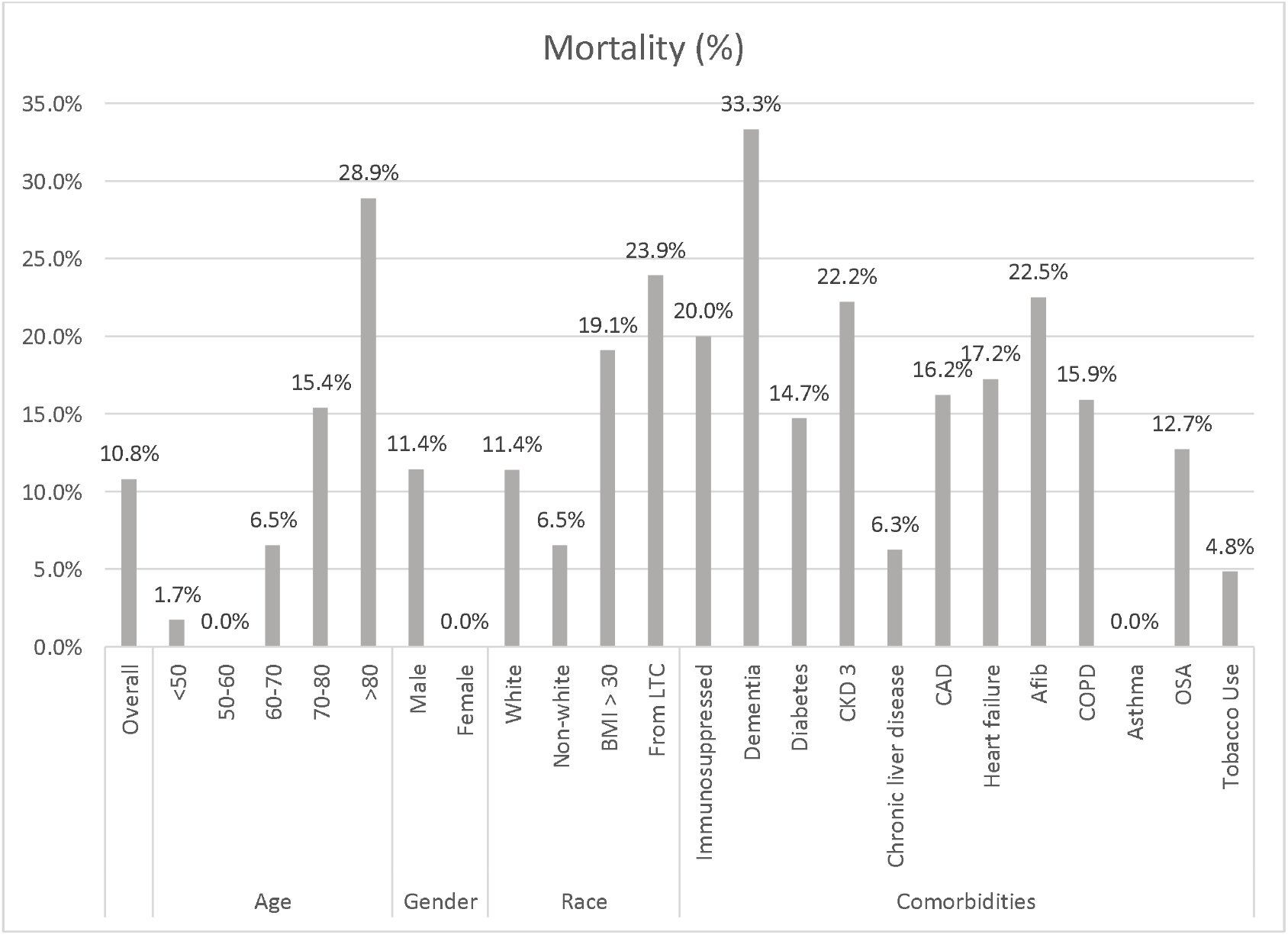
Percent of patients who died, based on patient demographics and comorbidities

